# Sociodemographic factors and self-restraint from social behaviors during the COVID-19 pandemic in Japan: a cross-sectional study

**DOI:** 10.1101/2021.12.27.21268446

**Authors:** Takahiro Mori, Tomohisa Nagata, Kazunori Ikegami, Ayako Hino, Seiichiro Tateishi, Mayumi Tsuji, Shinya Matsuda, Yoshihisa Fujino, Koji Mori, for the CORoNaWork project

**Author notes:** Correspondence author: Koji Mori, MD, PhD, Department of Occupational Health Practice and Management, Institute of Industrial Ecological Sciences, University of Occupational and Environmental Health, Japan, 1-1 Iseigaoka, Yahatanishi-ku, Kitakyushu 807-8555, Japan.

## Abstract

The control of human flow has led to better control of COVID-19 infections. Japan’s state of emergency, unlike other countries, is not legally binding but is rather a request for individual self-restraint; thus, factors must be identified that do not respond to self-restraint, and countermeasures considered for those factors to enhance its efficacy. We examined the relationship between sociodemographic factors and self-restraint toward going out in public during a pandemic in Japan. This cross-sectional study used data for February 18–19, 2021, obtained from an internet survey; 19,560 participants aged 20–65 were included in the analysis. We identified five relevant behaviors: (1) taking a day trip; (2) eating out with five people or more; (3) gathering with friends and colleagues; (4) shopping for other than daily necessities; (5) shopping for daily necessities. Multilevel logistic regression analyses were used to examine the association between sociodemographic factors and self-restraint for each of the behaviors. Results showed that for behaviors other than shopping for daily necessities, women, those aged 60–65, married people, highly educated people, high-income earners, desk workers and those who mainly work with interpersonal communication, and those with underlying disease reported more self-restraint. Older people had less self-restraint than younger people toward shopping for daily necessities; an underlying disease had no effect on the identified behavior. Specialized interventions for these groups that include recommendations for greater self-restraint may improve the efficacy of the implementing measures that request self-restraint.

## Introduction

Legally binding lockdown policies have been implemented in many countries in response to the spread of coronavirus disease 2019 (COVID-19) infections, which has led to the control of human flow and thus to improved control of infection transmission. In China, the first lockdown reduced inflows to the city of Wuhan by 76%, outflows by 56%, and movement within Wuhan by 54% [1]. In Italy, lockdown reduced interstate travel by 40% and the radius of action in daily life by 49% [2]; in France, the overall movement of people decreased by 65%, and the radius of action also decreased significantly [3]. Using longitudinal data from 202 countries, Aflano and Ercolano showed that controlling human flow via lockdowns reduced the number of newly infected people [4].

Japan’s state of emergency is not legally binding, unlike many other countries’ measures, but is rather a request for self-restraint. Therefore, infection control is left to the discretion of the individual. Japan’s state of emergency has been declared four times since the beginning of the COVID-19 pandemic: from April to May 2020, from January to March 2021, from April to June 2021, and from July to September 2021 [5]. When Japan’s government declares a state of emergency, the citizens are required to refrain from unnecessary and nonurgent social behaviors, including traveling and returning home, eating and drinking at a restaurant for extended periods of time, and socializing with groups of more than five people [6]. However, in the first state of emergency, when sufficient information about COVID-19 was not available, the government sought to forestall an emergency by prohibiting at least 70% of all people from going out of their homes [7]. As a result, movement across prefectures was suppressed by about 50% to 70% throughout Japan [8]. The public’s response to the second and subsequent states of emergency has gradually declined [8].

The impact of sociodemographic factors on social behaviors—specifically the act of leaving one’s home to visit public spaces—during lockdowns has been investigated mainly in Western countries. It has been reported that women [9-18] highly educated people, and high-income individuals [11,19-21] are more likely to refrain from going out during a lockdown. Whether a person is married or lives with their family [12,18,19], and whether or not they have a pre-existing condition [13,20], are reportedly not associated with social behaviors during a lockdown. Reports implicating age as a factor vary: many show that older people refrain from going out more than younger people [11-14,22], while others indicate the reverse [9,10].

During the first state of emergency in Japan, reports noted that women and young people, lower-income individuals, people living with two or more others, unemployed people, and those with a chronic disease were the least likely to leave their homes [23, 24]. However, to our knowledge, the relationship between sociodemographic factors and restraining oneself from social behaviors has not been investigated in Japan since the second and subsequent emergency declarations, when valid information on COVID-19 has been available and the public’s response to the states of emergency has diminished.

Especially when implementing measures that request self-restraint, it is necessary to identify factors that do not respond to self-restraint and to consider countermeasures for those factors to enhance the efficacy of those measures. Therefore, the purpose of this study was to examine the relationship between sociodemographic factors and the self-restraint of social behaviors during the declaration of the second state of emergency in Japan.

## Methods

### Study design and participants

We conducted a cross-sectional study using data from the Collaborative Online Research on Novel-coronavirus and Work study (CORoNaWork study) that was collected on February 18 and 19, 2021. This survey was conducted as a self-administrated questionnaire by the internet survey company, Cross Marketing Inc. (Tokyo, Japan). The CORoNaWork study started in December 2020 and recruited participants aged 20–65 years old who had employment contracts. Respondents to the CORoNaWork study were stratified by cluster sampling according to gender, age, and region of residence. Details of the study protocol have been previously reported [25]. Out of a total of 33,302 people who participated in December 2020, 19,560 were included in our analysis, after excluding non-responders, those who gave incorrect answers, and unemployed people at the time of our survey. The first state of emergency was declared in all prefectures, whereas the second and subsequent declarations were given only in those prefectures where the infection situation was worsening. Importantly, instances of citizens going out of their homes were considerably suppressed not only in those prefectures in which a state of emergency was declared, but also in prefectures in which no declaration was made [8,26]. Therefore, this survey targeted respondents from all prefectures.

The present study was approved by the Ethics Committee of the University of Occupational and Environmental Health, Japan (Approval numbers: R2-079 and R3-006). Informed consent was obtained in the form of the website from all participants.

### Assessment of social behaviors

We identified five distinct types of social behavior during a state of emergency; these reflect the Japanese government’s request for self-restraint [6]: (1) taking a day trip; (2) eating out with five people or more; (3) gathering with friends and colleagues; (4) shopping for other than daily necessities; (5) shopping for daily necessities. Although the government did not include shopping for daily necessities in their request for self-restraint [7], such places can be crowded, so we included it in this study to expand our analysis. For each of the five behaviors, we asked participants, “Has your self-restraint changed in response to the second declaration of a state of emergency in January 2021?” Respondents chose one of the following five options: “greatly increased,” “increased a little,” “no change,” “decreased a little,” or “greatly decreased.” We created a binary variable by defining “decreased a little” or “greatly decreased” as having self-restraint, and the other options as not having self-restraint.

### Assessment of sociodemographic factors

We investigated gender, age, education, marital status, household income, job type, and underlying disease. Age was classified into five groups: 20–29, 30–39, 40–49, 50–59, and 60–65 years old. Education was classified into three categories: junior high or high school, vocational school or college, and university or graduate school. Marital status was classified into three categories: married, divorced or widowed, and never married. Annual household income was classified into four groups: <4.00 million Japanese yen (JPY); 4.00–5.99 million JPY; 6.00–8.99 million JPY; and ≥9.00 million JPY (1 USD was equal to 106.78 JPY, using 2020 conversion rates) [27]. Job type was classified into four categories: mainly desk work, jobs mainly involving interpersonal communication, mainly physical work. Regarding underlying disease, we asked the question, “Do you have any disease that requires regular visits to the hospital or treatment?” Participants selected one of the following: “I do not have such a disease,” “I have such a disease,” or “I have such a disease, but refrained from going to the hospital following the second declaration of the state of emergency.” We rated the participants who answered, “I do not have such a disease” as “No” and the remaining two answers as “Yes.”

### Statistical analyses

Multilevel logistic regression analyses were used to examine the association between sociodemographic factors and self-restraint in each of the five identified social behaviors after the second declaration of a state of emergency. Age-gender adjusted odds ratios (ORs) were estimated with a multilevel logistic model nested in the prefecture of residence because of the influence of regional differences in the infection status of COVID-19. A p-value of less than 0.05 was considered statistically significant. All analyses were conducted using Stata Statistical Software (Release 16; StataCorp LLC, College Station, TX, USA).

## Results

Table 1 shows the characteristics of participants. A total of 19,560 people (10,978 men and 8,582 women) were analyzed. There were 6,465 people (33.1%) who had an underlying disease. The proportion of those who refrained from going out during the state of emergency were those who avoided: taking a day trip (11,661: 59.6%), eating out with five people or more (11,447: 58.5%), gathering with friends and colleagues (12,936: 66.1%), shopping for other than daily necessities (8,668: 44.3%), and shopping for daily necessities (5,392: 27.6%).

**Table 1.**
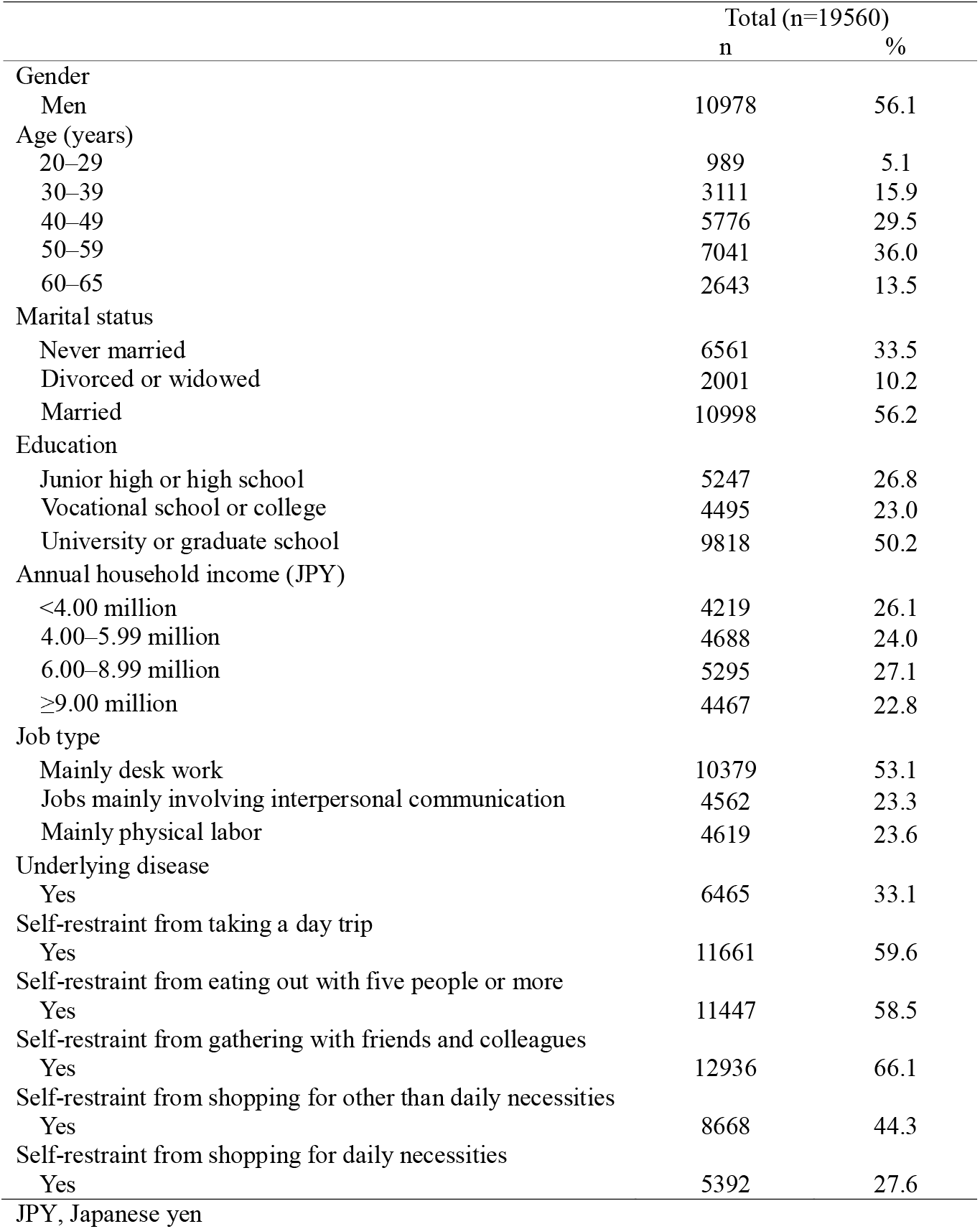
Characteristics of participants

|  | Total (n=19560) |  |
| --- | --- | --- |
|  | n | % |
| Gender |  |  |
| Men | 10978 | 56.1 |
| Age (years) |  |  |
| 20–29 | 989 | 5.1 |
| 30–39 | 3111 | 15.9 |
| 40–49 | 5776 | 29.5 |
| 50–59 | 7041 | 36.0 |
| 60–65 | 2643 | 13.5 |
| Marital status |  |  |
| Never married | 6561 | 33.5 |
| Divorced or widowed | 2001 | 10.2 |
| Married | 10998 | 56.2 |
| Education |  |  |
| Junior high or high school | 5247 | 26.8 |
| Vocational school or college | 4495 | 23.0 |
| University or graduate school | 9818 | 50.2 |
| Annual household income (JPY) |  |  |
| <4.00 million | 4219 | 26.1 |
| 4.00–5.99 million | 4688 | 24.0 |
| 6.00–8.99 million | 5295 | 27.1 |
| ≥9.00 million | 4467 | 22.8 |
| Job type |  |  |
| Mainly desk work | 10379 | 53.1 |
| Jobs mainly involving interpersonal communication | 4562 | 23.3 |
| Mainly physical labor | 4619 | 23.6 |
| Underlying disease |  |  |
| Yes | 6465 | 33.1 |
| Self-restraint from taking a day trip |  |  |
| Yes | 11661 | 59.6 |
| Self-restraint from eating out with five people or more |  |  |
| Yes | 11447 | 58.5 |
| Self-restraint from gathering with friends and colleagues |  |  |
| Yes | 12936 | 66.1 |
| Self-restraint from shopping for other than daily necessities |  |  |
| Yes | 8668 | 44.3 |
| Self-restraint from shopping for daily necessities |  |  |
| Yes | 5392 | 27.6 |
| JPY, Japanese yen |  |  |

Table 2 shows the sociodemographic factors associated with self-restraint from going out during a state of emergency. Women had higher odds ratios (ORs) than men for self-restraint from taking a day trip (OR 1.42), eating out with five people or more (OR 1.31), gathering with friends and colleagues (OR 1.55), shopping for other than daily necessities (OR 1.79), and shopping for daily necessities (OR 1.51). Compared with those aged 20–29, those aged 50–59 had a higher OR for self-restraint from gathering with friends and colleagues (OR 1.23), and those aged 60–65 had higher ORs for self-restraint from eating out with five people or more (OR 1.26), gathering with friends and colleagues (OR 1.61), and shopping for other than daily necessities (OR 1.18). However, regarding shopping for daily necessities, those aged 50–59 had a lower OR than those aged 20–29 (OR 0.83). Married people had higher ORs for self-restraint from all behaviors than people who had never been married. Highly educated people and high-income earners considerably refrained from going out except for shopping for daily necessities during the state of emergency. Compared with those whose job entailed mainly desk work, people with jobs mainly involving interpersonal communication had a higher OR for self-restraint from taking day trips (OR 1.09). Compared with desk workers, people whose jobs entailed mainly physical work had lower ORs for self-restraint from going out, except for shopping for daily necessities. People who had an underlying disease had higher ORs for self-restraint from going out, except for shopping for daily necessities.

**Table 2.** Relationship between sociodemographic factors and self-restraint from social behaviors during a state of emergency

|  | Self-restraint from taking a day trip |  |  | Self-restraint from eating out with five people or more |  |  | Self-restraint from gathering with friends and colleagues |  |  | Self-restraint from shopping for other than daily necessities |  |  | Self-restraint from shopping for daily necessities |  |  |
| --- | --- | --- | --- | --- | --- | --- | --- | --- | --- | --- | --- | --- | --- | --- | --- |
|  | OR* | 95% CI | *P* value | OR* | 95% CI | *P* value | OR* | 95% CI | *P* value | OR* | 95% CI | *P* value | OR* | 95% CI | *P* value |
| Gender |  |  |  |  |  |  |  |  |  |  |  |  |  |  |  |
| Men | 1.00 |  |  | 1.00 |  |  | 1.00 |  |  | 1.00 |  |  | 1.00 |  |  |
| Women | 1.42 | 1.34–1.52 | <0.001 | 1.31 | 1.23–1.39 | <0.001 | 1.55 | 1.45–1.65 | <0.001 | 1.79 | 1.69–1.91 | <0.001 | 1.51 | 1.41–1.61 | <0.001 |
| Age (years) |  |  |  |  |  |  |  |  |  |  |  |  |  |  |  |
| 20–29 | 1.00 |  |  | 1.00 |  |  | 1.00 |  |  | 1.00 |  |  | 1.00 |  |  |
| 30–39 | 1.01 | 0.87–1.18 | 0.855 | 0.98 | 0.85–1.14 | 0.822 | 1.06 | 0.91–1.24 | 0.427 | 1.07 | 0.93–1.24 | 0.359 | 0.97 | 0.83–1.13 | 0.679 |
| 40–49 | 0.95 | 0.82–1.09 | 0.471 | 0.97 | 0.84–1.11 | 0.637 | 1.11 | 0.96–1.28 | 0.163 | 1.06 | 0.92–1.22 | 0.406 | 0.92 | 0.79–1.06 | 0.250 |
| 50–59 | 0.93 | 0.81–1.08 | 0.337 | 1.00 | 0.87–1.15 | 1.000 | 1.23 | 1.06–1.42 | 0.006 | 1.03 | 0.89–1.18 | 0.700 | 0.83 | 0.72–0.96 | 0.013 |
| 60–65 | 1.16 | 0.99–1.36 | 0.063 | 1.26 | 1.08–1.47 | 0.004 | 1.61 | 1.37–1.90 | <0.001 | 1.18 | 1.01–1.38 | 0.035 | 0.85 | 0.72–1.01 | 0.063 |
| Marital status |  |  |  |  |  |  |  |  |  |  |  |  |  |  |  |
| Never married | 1.00 |  |  | 1.00 |  |  | 1.00 |  |  | 1.00 |  |  | 1.00 |  |  |
| Divorced or widowed | 1.12 | 1.01–1.24 | 0.037 | 1.14 | 1.02–1.26 | 0.015 | 1.14 | 1.02–1.27 | 0.020 | 0.96 | 0.86–1.07 | 0.444 | 1.09 | 0.97–1.22 | 0.164 |
| Married | 1.53 | 1.43–1.63 | <0.001 | 1.53 | 1.43–1.64 | <0.001 | 1.51 | 1.41–1.61 | <0.001 | 1.28 | 1.20–1.37 | <0.001 | 1.24 | 1.15–1.33 | <0.001 |
| Education |  |  |  |  |  |  |  |  |  |  |  |  |  |  |  |
| Junior high or high school | 1.00 |  |  | 1.00 |  |  | 1.00 |  |  | 1.00 |  |  | 1.00 |  |  |
| Vocational school or college | 1.36 | 1.25–1.47 | <0.001 | 1.37 | 1.26–1.49 | <0.001 | 1.36 | 1.25–1.48 | <0.001 | 1.26 | 1.16–1.36 | <0.001 | 1.07 | 0.98–1.17 | 0.153 |
| University or graduate school | 1.43 | 1.33–1.53 | <0.001 | 1.55 | 1.45–1.66 | <0.001 | 1.51 | 1.41–1.63 | <0.001 | 1.36 | 1.27–1.46 | <0.001 | 1.06 | 0.98–1.14 | 0.160 |
| Annually household income (JPY) |  |  |  |  |  |  |  |  |  |  |  |  |  |  |  |
| <4.00 million | 1.00 |  |  | 1.00 |  |  | 1.00 |  |  | 1.00 |  |  | 1.00 |  |  |
| 4.00–5.99 million | 1.30 | 1.20–1.41 | <0.001 | 1.30 | 1.20–1.41 | <0.001 | 1.37 | 1.26–1.49 | <0.001 | 1.21 | 1.12–1.32 | <0.001 | 1.14 | 1.04–1.25 | 0.004 |
| 6.00–8.99 million | 1.69 | 1.56–1.83 | <0.001 | 1.70 | 1.57–1.84 | <0.001 | 1.71 | 1.58–1.86 | <0.001 | 1.38 | 1.27–1.49 | <0.001 | 1.25 | 1.15–1.37 | <0.001 |
| ≥9.00 million | 1.85 | 1.70–2.01 | <0.001 | 1.99 | 1.83–2.16 | <0.001 | 1.95 | 1.78–2.13 | <0.001 | 1.45 | 1.33–1.58 | <0.001 | 1.19 | 1.09–1.31 | <0.001 |
| Job type |  |  |  |  |  |  |  |  |  |  |  |  |  |  |  |
| Mainly desk work | 1.00 |  |  | 1.00 |  |  | 1.00 |  |  | 1.00 |  |  | 1.00 |  |  |
| Jobs mainly involving interpersonal communication | 1.09 | 1.01–1.17 | 0.023 | 1.04 | 0.96–1.11 | 0.325 | 1.06 | 0.99–1.15 | 0.107 | 0.99 | 0.92–1.07 | 0.829 | 0.97 | 0.90–1.05 | 0.496 |
| Mainly physical labor | 0.80 | 0.74–0.85 | <0.001 | 0.71 | 0.66–0.76 | <0.001 | 0.75 | 0.70–0.81 | <0.001 | 0.83 | 0.77–0.89 | <0.001 | 0.96 | 0.88–1.04 | 0.284 |
| Underlying disease |  |  |  |  |  |  |  |  |  |  |  |  |  |  |  |
| No | 1.00 |  |  | 1.00 |  |  | 1.00 |  |  | 1.00 |  |  | 1.00 |  |  |
| Yes | 1.13 | 1.06–1.20 | <0.001 | 1.13 | 1.06–1.20 | <0.001 | 1.15 | 1.07–1.22 | <0.001 | 1.07 | 1.01–1.14 | 0.024 | 0.98 | 0.91–1.05 | 0.497 |

\*Adjusted for age and gender
JPY, Japanese yen

## Discussion

We investigated the association between sociodemographic factors and self-restraint from social behaviors after the second state of emergency was declared in Japan. The results showed that the most self-restraint was reported with respect to going out aside from shopping for daily necessities, which are regarded as unnecessary and nonurgent social behaviors, by the following groups: women, those aged 60–65, married people, highly educated people, high-income earners, desk workers and those who mainly work with interpersonal communication, and those with an underlying disease. By contrast, women, married people, and high-income earners reported substantial self-restraint from shopping for daily necessities (as much as that for other types of outings); however, those aged 50-59 had lower self-restraint than those aged 20-29, and those aged 60-65 did not show a significant difference, but there was a tendency for less self-restraint. No difference was observed for educational background, job type, or presence of underlying disease. Our findings suggest that it is necessary to consider measures to increase individuals’ self-restraint during states of emergency toward both unnecessary and nonurgent social behaviors and shopping for daily necessities which is unavoidable to some extent.

Our findings were consistent with those produced during Japan’s first state of emergency; specifically, that women, married people, people living with others, and those with a chronic disease refrained the most from going out. However, our results differed in terms of age and income class [23,24]. Our study was conducted approximately one year after a COVID-19 infection was first reported in Japan [28], and since then, a lot more information on COVID-19 has been accumulated. Thus, it was possible for people to judge, on the basis of that information, whether to refrain from social behaviors, a situation unlike the investigations conducted during the first state of emergency [23,24].

Our findings are also consistent with other countries’ reports with respect to gender, age, educational background, and income, but differ slightly with respect to marriage and underlying diseases. According to the COVID-19 Stringency Index published by Oxford University, which measures and indexes the strength of each country’s infection control, Japan ranks considerably lower than China, European countries, and the United States, all of which have implemented lockdown measures [29]. Japan’s state of emergency is not legally binding, so it is up to the individual to respond to requests for self-restraint, a factor that may help explain the differences between Japan’s and other countries’ in some aspects of their respective reports.

Self-restraint from social behaviors is influenced by the perception of the risk regarding infectious diseases [30-32], including an awareness of the possibility and severity of infection [31]. More specifically, it has been reported that awareness of potential severity easily leads to self-restraint in socializing behaviors [33,34]. Concerns about not only one’s own health but also the health of family and friends reportedly affect this self-restraint [15]. Therefore, it is conceivable that someone will refrain from going places with high risk of infection in consideration of the possibility that they will be infected and then transmit their infection to a family member or acquaintance.

### Unnecessary and nonurgent social behaviors

One of the reasons women refrained from going out during the state of emergency more than men was likely the influence of risk perception. It is reported that women have a higher risk perception for various hazards, including during the SARS outbreak [35,36]. Furthermore, women are more likely to actively collect information on COVID-19 [37], and generally spend more time raising and caring for children than men, especially during the COVID-19 pandemic [38]. Therefore, they may be more aware of their impact on others and thus concerned not only about being infected themselves but also about their children and the elderly people they care for being infected, which may increase their risk-avoidance.

The fact that those aged 60–65 years old refrained from going out is arguably also an indication of this group’s risk perception, given the data that show elderly people are more likely to become severely ill if they contract COVID-19 [39]. Indeed, the government and the media reported on this daily, emphasizing that many serious illnesses and deaths were experienced largely by elderly people, which may have contributed to the increased risk perception among the elderly population that led to self-restraint. For example, the substantial self-restraint in this group toward eating out with five or more people and gathering with friends and colleagues, may reflect their concern about being infected and transmitting the virus to their peers, who might then become seriously ill.

Other groups who evidenced higher risk perception (in the form of self-restraint) were: married people, highly educated and high-income earners, desk workers and those who mainly work with interpersonal communication, and those with an underlying disease. Married people are likely aware of their domestic situation as an infection route [40,41] and thus pay more attention to the possible ways in which their family members may become infected, in addition to their own infection risk, which may have contributed to their self-restraint. The self-restraint behaviors of the highly educated and the high-income earners may be explained by their generally high health literacy [43,44], which conceivably stems from their ability and desire to actively collect and select valid information, resulting in increased risk perception and the adoption of behaviors that protect them from COVID-19 [34].For many people whose jobs involve mainly desk work, they are often able to work from home, which in itself is part of the recommended infection control measures and may therefore raise these workers’ risk perception. Even in workplaces where people engage in interpersonal communication, measures against infection have been strengthened to avoid infecting customers as well as employees. One report showed that infection control measures in the workplace cause individuals to strengthen their own protective measures [44]; thus, such workplace measures may naturally raise an individual worker’s risk perception. Finally, people who have an underlying disease are at an increased risk of becoming more severely ill if they contract COVID-19 [39], information that is, at least in Japan, repeatedly disseminated by the government and experts, which may have raised the perception of risk in this group and led to the reported self-restraint.

### Shopping for daily necessities

Our results showed that, in contrast to younger people, older people did not refrain from shopping for daily necessities, and we observed no difference in this behavior between those with and those without an underlying disease; this was unlike all other self-restraint scenarios in this study. One of the reasons for this may have been that they are not as aware of the risk of getting infected while shopping as other social behaviors. One study reported that young people adhered more closely to the set shopping time [15], which indicates that young people may be more aware that shopping areas can be crowded and thus the risk of infection may be high. Another reason is that older people are less likely to use online shopping for those items or to use a food delivery service [45]. Furthermore, it is possible that shopping provided a community place for those older participants.

Women, married people, and high-income earners may be more aware of the risk of infection associated with shopping and with public outings that are unnecessary and nonurgent. Furthermore, high-income groups and people living with family were more likely to shop online [15,45], which helps explain how they can refrain from going out to shop.

### Implementation

During states of emergency in Japan, interventions to improve self-restraint behaviors to the groups lacking self-restraint toward unnecessary and nonurgent outings, such as men, young to middle-aged people, the less-educated, and those who do mainly physical work, are required. More specifically, interventions should draw on reports showing that trust in government and media influences risk perception [46], and in particular, information about the infection situation, including measures by the government, daily reports on the number of infected people, and deaths, has a great impact on self-restraint behaviors [26]. Therefore, it is important to take the group characteristics into account when considering the content and distribution channels of the information that is shared with the public.

For example, men are at higher risk of becoming more severely ill with COVID-19 than women [47,48], and those aged 40–59 are at higher risk of becoming more severely ill than younger people [39]; thus it is necessary, when creating content, to actively communicate these specific risks for certain groups of becoming more severely ill. To communicate to people with less education and lower incomes, it is important to account for their statistically lower literacy rates and send out simple, easy-to-understand information. Furthermore, because so much incorrect information and so many rumors continue to circulate, it is important to show how to identify correct information and thus avoid hindering effective responses and creating confusion and mistrust [49].

As examples of the routes to be considered for the dissemination of information, middle-aged and older people tend to obtain and trust information about COVID-19 from television and newspapers, while younger people tend to obtain and trust information from the Internet and social networking services [37]. For people who mainly do physical work, it is important to actively disseminate information in the workplace, because it is difficult to convey the importance of infection control through work styles.

With respect to shopping for daily necessities, we think it is necessary to recommend online shopping for elderly people and for people with an underlying disease, and to consider service methods that can be easily used even by people who are not familiar with information technology. We also think that it is necessary to disseminate information that encourages healthy people to do the necessary shopping rather than those whose health is compromised. However, because shopping for daily necessities entails an unavoidable outing to some extent, finding ways to avoid crowded places is also important; this could be accomplished by dividing shopping time by age, limiting the time spent shopping, and limiting the number of people who go shopping.

### Limitations

This study had some limitations. First, we conducted an internet survey, which includes the possibility of selection bias. To reduce the potential bias as much as possible, sampling was balanced by gender, age, occupation, and area of residence. Second, the outcome variable was self-restraint from social behaviors, which was measured via subjective evaluations; moreover, the specific degree of self-restraint was not investigated. When the second state of emergency was declared in Japan, there was no specific target value for such self-restraint behaviors from the government, so we investigated whether self-restraint changed in response to state of emergency Third, this study was cross-sectional in nature. Our results are different from the survey that was conducted during the first state of emergency in Japan [23,24], so it is possible that the results will continue to change over time. Further investigation is required to see if the same results are obtained for the time period covering the third and subsequent state of emergency.

## Conclusions

Self-restraint from social behaviors during a pandemic is an important concept for developing measures to prevent the spread of infectious diseases. Our results showed that the groups who had the least self-restraint in unnecessary and nonurgent contexts were men, young to middle-aged people, people who had never been married, people with less education, people with lower incomes, and people whose work was mainly physical. Regarding shopping for daily necessities, older people had low self-restraint. Interventions designed specifically for these groups that recommend self-restraint from social behaviors may enhance the efficacy of the implementing measures that request self-restraint.

## Data Availability

Data not available due to ethical restrictions

## Abbreviations

COVID-19: Coronavirus disease 2019
CORoNaWork: Collaborative Online Research on the Novel-coronavirus and Work
JPY: Japanese yen
OR: Odds ratio

## Declarations

## Acknowledgments

The current members of the CORoNaWork Project, in alphabetical order, are as follows: Dr. Yoshihisa Fujino (present chairperson of the study group), Dr. Akira Ogami, Dr. Arisa Harada, Dr. Ayako Hino, Dr. Hajime Ando, Dr. Hisashi Eguchi, Dr. Kazunori Ikegami, Dr. Kei Tokutsu, Dr. Keiji Muramatsu, Dr. Koji Mori, Dr. Kosuke Mafune, Dr. Kyoko Kitagawa, Dr. Masako Nagata, Dr. Mayumi Tsuji, Ms. Ning Liu, Dr. Rie Tanaka, Dr. Ryutaro Matsugaki, Dr. Seiichiro Tateishi, Dr. Shinya Matsuda, Dr. Tomohiro Ishimaru, and Dr. Tomohisa Nagata. All members are affiliated with the University of Occupational and Environmental Health, Japan.

